# Post-peak mpox in England: epidemiology, reinfection, and vaccine effectiveness – data from 2023

**DOI:** 10.1101/2024.02.26.24303362

**Authors:** Hannah Charles, Katie Thorley, Charlie Turner, Kirsty F. Bennet, Nick Andrews, Marta Bertran, Sema Mandal, Gayatri Amirthalingam, Mary E. Ramsay, Hamish Mohammed, Katy Sinka

## Abstract

England, like other countries that experienced a large outbreak of emergent mpox in 2022, continued to record cases during 2023 at a low but steady frequency. Comprehensive national surveillance shows that cases continue to occur primarily among gay, bisexual and other men who have sex with men mostly in London. Of 137 cases in 2023, around half were acquired overseas, half were vaccinated, and one case of reinfection was reported. Eleven people required hospital care. High vaccination uptake, during 2022, has provided good coverage of those at higher risk of mpox. Using the screening method, vaccine effectiveness of one dose was estimated at 84%. None of the vaccinated cases in 2023 were hospitalised.

## Introduction

In May 2022, an international outbreak of mpox was identified, primarily affecting highly interconnected sexual networks of gay, bisexual and other men who have sex with men (GBMSM). In England, the number of mpox cases peaked in mid-July 2022 at 350 cases per week (1), and then declined to fewer than six cases per week from November onwards (2). From June 2022, the Modified Vaccinia Ankara – Bavarian Nordic (MVA-BN) vaccine was offered to GBMSM in England who were assessed as being at higher risk of mpox infection, including those with a recent history of multiple partners, participating in group sex or attending sex-on-premises venues (3), with the estimated vaccine effectiveness of one dose being 78% (4). Although the number of cases declined, mpox cases continued to be reported throughout 2023. Surveillance has continued to detect changes in the epidemiology and record risk factors that contribute to the ongoing occurrence of new mpox cases. This analysis summarises the epidemiology of post-peak mpox cases in 2023 in England (01 January to 31 December), describes case characteristics including vaccination status, investigates if characteristics varied to those from the peak of the outbreak, and estimates the effectiveness of vaccination to protect against symptomatic disease.

## Materials and Methods

Records of confirmed and highly probable mpox diagnoses reported between 01 January and 31 December 2023 were extracted from the UK Health Security Agency (UKHSA) Second Generation Surveillance System (SGSS) and deduplicated using specimen and patient identifiers. A confirmed case was defined as a positive *monkeypox virus*-specific polymerase chain reaction (PCR) and a highly probable case as a positive *Orthopoxvirus* PCR (5). SGSS receives positive test results from all diagnostic laboratories in England (6). Given the initial status of mpox as a high consequence infectious disease (HCID) and ongoing status as a statutory notifiable disease (7), in addition to the World Health Organization (WHO) classifying the 2022 outbreak as a Public Health Emergency of International Concern (PHEIC) (8), the reporting of mpox diagnoses to SGSS is very likely to be complete.

Additional epidemiological and behavioural information was collected by UKHSA’s local Health Protection Teams. Aggregate data on mpox vaccinations administered in 2022 and 2023 were provided by National Health Service (NHS) England. Overall vaccine effectiveness was estimated using the screening method (9). Difference between one dose and two dose vaccine effectiveness estimates were tested by a chi-squared test at a 5% significance level.

## Results

There were 137 mpox cases in the period between 01 January and 31 December 2023, 135 were confirmed cases and two were highly probable. Following an initial period of low-level reporting, with nine cases in total between January and March (Week 1 to Week 13) 2023, there was an increase in the frequency of cases, with a further 128 cases between April and the end of December (Week 17 to Week 52) (Figure 1). Consistent with the outbreak overall, of the 137 mpox cases in 2023, the majority (105 out of 137; 77%) were resident in London; the remaining 32 cases were reported from the East Midlands, South East, Yorkshire and Humber, East of England, North West and South West of England.

**Figure 1.**
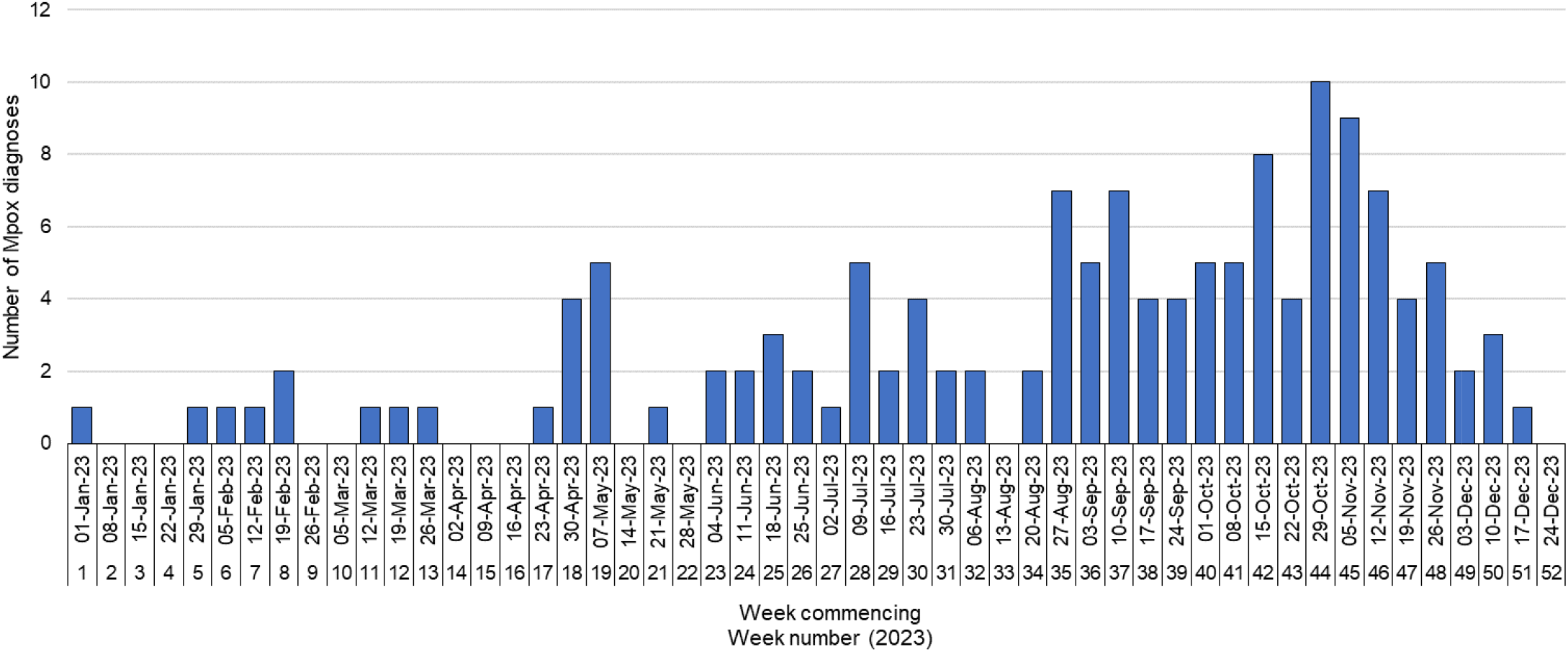
Confirmed and highly probable mpox cases by week of specimen date in England, 01 January to 31 December 2023.

Most cases identified as GBMSM (107 out of 137; 78%), an additional 21 cases were adult men without recorded information on sexual orientation and for this analysis of cases were assumed to be part of the 2022-23 global mpox outbreak, totalling 128 (out of 137; 93%). Eight cases were not considered to be part of this outbreak given information linking their infection to an mpox endemic country, these were sporadic cases distributed across the time period and consisted of heterosexual individuals and children. There was one further adult case where neither gender nor sexual orientation was reported.

Of the 137 cases in 2023, 64 individuals (47%) reported they did not travel outside the United Kingdom (UK) in the 21 days prior to symptom onset, indicating that they likely acquired mpox in the UK; 57 (89%) of these individuals identified as GBMSM (Table 1).

**Table 1:** Exposure classification of confirmed and highly probable mpox cases, by gender and sexual orientation, England, 01 January to 31 December 2023.

|  | Cases presumed acquired in the UK | Cases presumed acquired outside the UK | Country of acquisition unknown |
| --- | --- | --- | --- |
| Gender and sexual orientation | n (%)* | n (%)* | n (%)* |
| GBMSM | 57 (89) | 43 (81) | 6 (30) |
| Non-GBMSM or unknown | 7 (11) | 10 (19) | 14 (70) |
| **Total** | **64** | **53** | **20** |

GBMSM: Gay, bisexual and other men who have sex with men
\* Percentages are column percentages

Fifty-three individuals (39%) reported travel outside the UK, and of these, 43 (81%) identified as GBMSM and reported travelling to countries in Europe, the Middle East, Asia Pacific and North America.

Over half of cases in 2023 associated with the 2022-23 global mpox outbreak were HIV negative (69 out of 128; 54%), of whom most (51 out of 69; 74%) were taking HIV pre-exposure prophylaxis (PrEP). Thirty-nine individuals (39 out of 128; 30%) reported attending an event involving sexual contact with multiple partners and 24 individuals (19%) were diagnosed with a concurrent STI. One individual reported also having a previous mpox diagnosis in 2022 (Table 2).

**Table 2:** Characteristics and risk factors of confirmed and highly probable mpox cases associated with the 2022-23 global mpox outbreak, England, 01 January to 31 December 2023.

| **HIV status** | **n (%)** |
| --- | --- |
| Living with HIV | 13 (10) |
| HIV negative | 69 (54) |
| Taking HIV PrEP* | 51 (74) |
| Not taking PrEP* | 18 (26) |
| Unknown | 46 (36) |
| **Vaccination status** |  |
| Vaccinated – $\geq 2$ doses ☐ | 30 (23) |
| Vaccinated – 1 dose | 20 (16) |
| Vaccinated – number of doses unknown | 2 (2) |
| Unvaccinated | 54 (42) |
| Unknown | 22 (17) |
| **Hospital admission** |  |
| Yes | 11 (9) |
| No/unknown | 117 (91) |
| **Reporting attending events involving multiple sexual partners** |  |
| Yes | 39 (30) |
| No/unknown | 98 (70) |
| **Concurrent STI** |  |
| Yes | 24 (19) |
| No/unknown | 104 (81) |
| **Previous mpox diagnosis** |  |
| Yes | 1 (1) |
| No | 64 (50) |
| Unknown | 63 (49) |

HIV PrEP: HIV pre-exposure prophylaxis
Unless otherwise stated, the denominator is 128
\* Denominator for percentage calculations is HIV negative individuals
☐ Includes 1 individual who reported receiving three doses

Of cases associated with the 2022-23 global mpox outbreak, 52 (49%) of the 106 cases with known vaccination status were fully or partially vaccinated, of whom one case reported three doses, 29 cases received two doses and 20 cases received one dose, and 2 did not report the number of doses. Among these vaccinated cases, 20 (38%) reported attending an event involving sexual contact with multiple partners and were therefore likely to have been at higher risk of exposure to mpox. Eleven individuals were admitted to hospital due to their mpox infection (9%), nine were unvaccinated and two had unknown vaccination status.

### Overall vaccine effectiveness

Between July 2022 and December 2023, 77,543 GBMSM in England were vaccinated against mpox; 32,983 individuals with one dose and 44,560 with two. Based on an estimated eligible GBMSM denominator of 89,240 as used previously (4) this gives a coverage estimate at the end of December 2023 of 37.0% for one dose and 49.9% for two doses (86.9% one or two doses). Most vaccines had been given by March 2023 (91.0% of first doses and 75.7% of second doses).

Vaccine effectiveness was calculated using the screening method, as previously described (4). For the primary analysis, the eligible GBMSM denominator was taken to be 89,240 and 20% higher at 107,088 in a sensitivity analysis, similar to previous analyses (4). For the calculation of vaccine effectiveness, the proportion of GBMSM vaccinated was estimated based on matching each case to the one and two dose coverage at the time two weeks before individuals became a case, then averaging this matched coverage across cases. For the 22 cases where vaccination status was unknown, it was assumed that these would be distributed among those with 0, 1 and 2 doses in the same ratio as the observed ratio for these groups, to give a corrected value.

Vaccine effectiveness of a single dose was estimated at 84% (Table 3) - comparable to the previous estimate of 78% (4). The vaccine effectiveness of two doses was similar to one dose, at 80% (p=0.40). The overall vaccine effectiveness of one or two doses was 82%.

**Table 3:** Vaccination status of mpox cases during 2023 used to estimate vaccine effectiveness of one dose, two doses and one or two doses of Modified Vaccinia Ankara – Bavarian Nordic (MVA-BN) vaccine using the screening method.

| Doses | 2023 Cases | Corrected Cases* | PCV | PPV | PPV (sensitivity**) | VE (95% CI) | VE (95% CI) (sensitivity**) |
| --- | --- | --- | --- | --- | --- | --- | --- |
| 0 | 54 | 65.4 | 52% | 16% | 30% |  |  |
| 1 | 20 | 24.2 | 19% | 38% | 32% | 84% (74-91) | 65% (42-79) |
| 2 | 30 | 36.4 | 29% | 45% | 38% | 80% (69-83) | 56% (31-72) |
| Unknown status | 22 |  |  |  |  |  |  |
| 1 or 2 | 50 | 60.6 | 48% | 84% | 70% | 82% (74-88) | 60% (41-73) |
| Total | 126 | 126 |  |  |  |  |  |

PCV = proportion of cases with the given number of doses; PPV = proportion of the population with the given number of doses obtained by matching each case to the population uptake 14 days prior to onset and averaging this across cases; VE = vaccine effectiveness.
\* 22 cases with unknown vaccination status distributed among 0, 1 and 2 doses. \*\* based on the higher estimated GBMSM denominator (107,088)

None of the vaccinated cases in 2023 were hospitalised, compared to eleven among those unvaccinated (or where vaccination status was not known). The sensitivity analysis resulted in vaccine effectiveness estimates which were markedly lower, demonstrating that the method is affected by assumptions of the size of the GBMSM population.

## Discussion

The very low case numbers at the start of the 2023 were initially interpreted as the final few cases in the tail of the outbreak from 2022 (1). However, like other countries where the 2022 outbreak had been substantial, cases continued to occur steadily throughout the year with an even split of imported infections and community transmission.

The demographic and behavioural characteristics of mpox cases in 2023 remained similar to the peak of the outbreak in 2022 (10-12) and indicate that mpox continues to circulate predominately within interconnected sexual networks of GBMSM.

Nearly half of the cases in England in 2023 had been vaccinated and there were more cases among those who had received two doses of MVA-BN vaccine, compared to one dose. The vaccine effectiveness, influenced by the size of the GBMSM population, was similar for one dose and two doses, with the point estimate a little higher for one dose. Considering that first doses will have been given longer ago than second doses and that two dose would be expected to confer more protection the similar VE is counter intuitive. The similar VE may reflect differences in risk behaviour in those who came forward for a second dose of vaccine as they may also be more likely to come into contact with the monkeypox virus. This phenomenon of risk compensation has been suggested with other emerging infectious diseases such as COVID-19, particularly if behaviour modification associated with vaccination status occurs (13)(14). Increased awareness of the effectiveness of the MVA-BN vaccine and associated reduction in fear of mpox since the rollout of the vaccination programme may have also played a part. Critically, the finding that there were no hospitalised cases in 2023 among vaccinated individuals provides assurance that the MVA-BN vaccine protects against severe disease, which is corroborated by a global case series which found that illness among vaccinated individuals was less severe (15), and recent data from France (16).

The picture seen in England is consistent with reports from other high-income countries with outbreaks predominantly among GBMSM - in March 2023, reports of a cluster of mpox in France, included more vaccinated than unvaccinated individuals and in May 2023 the Chicago Department of Public Health noted that the majority of the cases reported since mid-April were among men who had received two doses of MVA-BN vaccine (17), yet a higher number of first doses had been given compared to second doses overall (18).

The self-reported reinfection in 2023 follows a previous case reported from 2022 (19). This finding, alongside the observed vaccine breakthrough infections, contributes to the emerging understanding of natural and vaccine induced immunity to mpox. Although the attending sexual health clinic verified that this individual had a positive mpox test in 2022 and another in 2023, it should be acknowledged that a lack of sequencing data cannot rule out persistent infection.

The 2023 overall experience in England was of continued low level community transmission among GBMSM, as well as imported infections, which looks set to continue. With over 20 countries continuing to report mpox cases in December 2023 and the WHO assessment that the overall global risk for men who have sex with men remains moderate (20), a forecast long tail to the outbreak is likely before elimination can be achieved. This underpins the importance of continuing with active prevention through vaccination and health promotion to those at higher risk, and ongoing surveillance to better understand factors that contribute to continued transmission.

## Data Availability

All data produced in the present work are contained in the manuscript

## Ethical statement

This analysis was undertaken for health protection purposes under permissions granted to UKHSA to collect and process confidential patient data under Regulation 3 of The Health Service (Control of Patient Information) Regulations 2020 and Section 251 of the National Health Service Act 2006.

## Acknowledgements

We would like to thank NHS England for their collection and sharing of vaccination data for the mpox vaccination programme and sexual health services for delivering the vaccines. We would also like to thank the UKHSA Health Protection Teams for collecting epidemiological information during public health management of cases.

## Authors’ contributions

HC led the epidemiological analysis and drafted the manuscript. The epidemiological data analysis was validated by KT and KFB. NA led the vaccine effectiveness analysis. All co-authors contributed to interpretation of the findings and to revision of the manuscript.

